# The UCLA ATLAS Community Health Initiative: promoting precision health research in a diverse biobank

**DOI:** 10.1101/2022.02.12.22270895

**Authors:** Ruth Johnson, Yi Ding, Arjun Bhattacharya, Alec Chiu, Clara Lajonchere, Daniel H. Geschwind, Bogdan Pasaniuc

## Abstract

The UCLA ATLAS Community Health Initiative (ATLAS) has an initial target to recruit 150,000 participants from across the UCLA Health system, with the goal of creating a genomic database to accelerate precision medicine efforts in California. This initiative includes a biobank embedded within the UCLA Health system that comprises de-identified genomic data linked to electronic health records (EHR). The first freeze of data from September 2020 contains 27,987 genotyped samples imputed to 7.9 million SNPs across the genome and is linked with a de-identified EHR extract. This database enables the study of numerous clinically-related phenotypes within the same medical system. Here we describe a centralized repository of the genotype data and provide tools and pipelines to perform genome-wide and phenome-wide association studies across a wide range of EHR-derived phenotypes and genetic ancestry groups. We demonstrate the utility of this resource through the analysis of 7 well-studied traits and recapitulate many previous genetic and phenotypic associations.

## The UCLA ATLAS Community Health Initiative

The UCLA ATLAS Community Health Initiative (ATLAS) aims to recruit 150,000 participants from across the UCLA Health system, with the goal of creating California’s largest genomic resource for translational and precision medicine research. Each biosample is linked with the patient’s electronic health record from UCLA Health via the UCLA Data Discovery Repository (DDR), a database containing a de-identified extract of electronic health records (EHR). Participants are recruited from 18 UCLA Health medical centers, laboratories, and clinics located throughout the greater Los Angeles area. Participants watch a short video outlining the goals of the initiative and document their choice of whether they wish to consent to participation [1,2].

Biological samples are collected during routine clinical lab work performed at any UCLA Health laboratory and then genotyped using the Illumina Global Screening Array (GSA) [3]. Both biological samples and EHR information are de-identified to protect patient privacy. As of September 2021, the initiative has enrolled 90,400 participants through the consent process and successfully genotyped 39,300 samples. Comprehensive details on the biobanking and consenting processes are described in prior work [1,2]. In this work, we describe quality control pipelines for genotype curation and phenotype extraction from the medical records for the purpose of large-scale genotype/phenotype scans. To establish the genotyping QC pipelines, we present the first freeze of the data containing genotypes/phenotypes collected and processed up to September 2020, resulting in a total of N=27,987 samples.

The UCLA Health System includes 2 hospitals and a total of 210 primary and specialty outpatient locations located primarily in the greater Los Angeles area. In total, the UCLA Health System serves approximately 5% of Los Angeles County. An electronic form of health records was implemented throughout the UCLA Health system in 2013, where a variety of clinical information is recorded, such as laboratory tests, medications and prescriptions, diagnoses, and hospital admissions. An extract of this information has been de-identified and approved for research purposes. The de-identification process removes some clinical data including names, family relationships, geographic information, exact dates, and exact ages for those at the extremes of age (>90 years old).

The average age of participants, defined as a participant’s age recorded in the EHR as of September 2021, is 55.6 (SD: 17.2) years with an average medical record length of 11.6 (SD: 8.5) years. We use phecodes, a coding system that maps diagnosis codes (e.g. ICD-9 and ICD-10 codes) to more clinically meaningful phenotypes [4], to construct phenotypes from the EHR. The median number of phecodes per participant is 68 whereas the mean is 85.2 (SD: 65.0). This skewed mean is consistent with the presence of individuals with many more healthcare interactions than the average person in the general population, a pattern that has been well described in the literature [5].

Participants’ self-identified race and ethnicity information is also recorded within the DDR where participants select a single option for their race and a single separate option for their ethnicity from provided multiple-choice lists. The majority of patients self-identify as White/Caucasian race (61.4%) and Non-Hispanic/Latino ethnicity (75.4%), although a substantial proportion of individuals report being of an Asian race (9.67%) or of Hispanic/Latino, Spanish, or Mexican ethnicity (14.1%). A full list of the provided race/ethnicity fields within the DDR and a summary of the ATLAS demographic information can be found in Table 1.

We regret that the term ‘White/Caucasian’ is a preset multiple-choice option under the race field within the medical records. For this reason, we choose to report subsequent analyses with this same language since the results should be referenced with the same terminology in which the data was collected (i.e. patients filled out medical forms with this specific phrasing). The scientific and medical communities have since denounced this specific terminology due to its erroneous origins and historically racist implications [6–8], but it is still built into the language of many documents and surveys, such as those within electronic health record systems. In presenting our analyses, we strongly discourage the connection of the term ‘Caucasian’ with the discussion of race, a social construct separate from biology, and emphasize that the term does not have any biological implications.

### Genotype generation and quality control

Genotyping was performed at the UCLA Neuroscience Genomics Core using a custom genotyping array constructed from the Global Screening Array with the multi-disease drop-in panel [3] under the GRCh37 assembly. An additional set of “Pathogenic” and “Likely Pathogenic” variants selected from ClinVar [9], such as the key SNPs found in the ACMG 59 genes [10], were also included in the chip design. Overall, the array measures 700,079 sites for capturing single nucleotide polymorphisms (SNPs) and short insertions and deletions (indels). Additional details regarding the array design are available on the Global Screening Array + Multi Disease SNP UCLA browser (https://coppolalab.ucla.edu/gclabapps/ungc/home).

The ATLAS initiative continuously recruits new participants and batches of genotype samples are being processed in monthly installments of approximately 1,000 samples per batch. The first freeze of genotype data presented in this work combines samples from 15 separate batches yielding a total of 697,023 SNPs and 27,987 individuals. Principal component analysis (PCA) [11] was used to visualize the variation across batches and did not identify any evidence of batch effects (Supplementary Figure S1).

We next describe the quality control pipeline used to filter out low-quality SNPs and samples while also considering the diverse ancestral backgrounds represented in ATLAS. First, we excluded poor-quality SNPs with >5% missingness as well as strand ambiguous and monomorphic SNPs. Samples with >5% missingness were also removed. We estimated kinship coefficients using KING 2.2.2 [12] and found 38 duplicate pairs of individuals (or twins), 357 parent-offspring, 128 first-degree, and 166 second-degree relatives. For the pairs of duplicate samples, we removed the sample with the higher missing rate. This level of relatedness is not surprising since members within a family tend to attend the same health center. A summary of the quality control pipeline and the number of filtered SNPs and individuals is outlined in Figure 1. Following sample- and variant-level quality control, M=673,130 genotyped SNPs remained across N=27,946 individuals (N=27,291 unrelated individuals).

**Figure 1:**
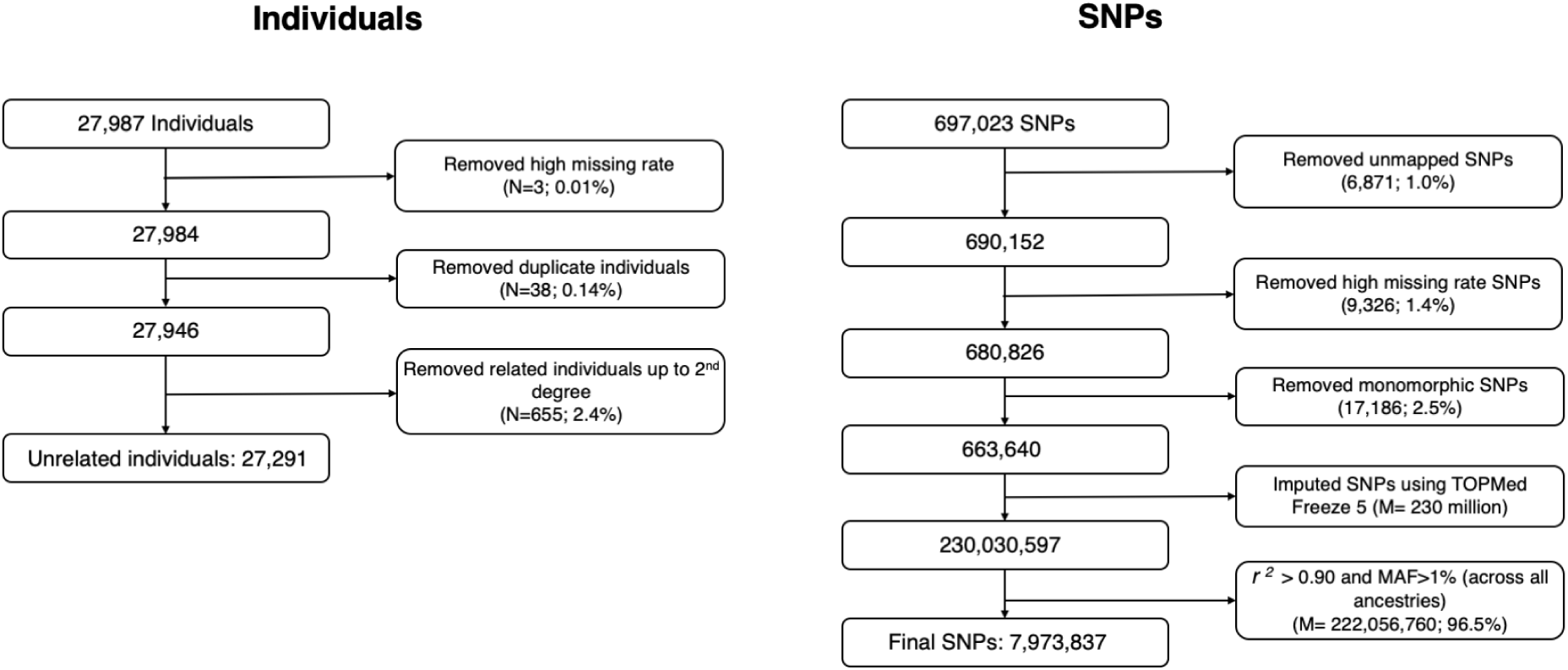
Summary of genotype quality control pipeline. We outline the quality control pipeline for the genotype samples and list the number of excluded samples (left) and SNPs (right) at each step.

After genotyping QC, we inferred biological sex using the ‘--sex-check’ function with default thresholds implemented in PLINK 1.9 [13] which estimates the X chromosome homozygosity or F statistic. We find that 45.5% of genotypes yield a male call and 53.9% a female call while 0.6% of samples were estimated to be unknown (Table 1). Using self-identified information from the EHR, we find that 45.1% of individuals self-identify as male and 54.9% self-identify as female (Table 1). Within the EHR, this specific field is labeled as ‘Sex’ and has a list of pre-determined multiple-choice fields where participants select one of the following options: ‘Male’, ‘Female’, ‘Other’, ‘Unknown’, ‘*Unspecified’, ‘X’. We observe that 0.04% of individuals who are inferred to be biologically male do not self-identify as male as reported from the EHR. This comparison is a common heuristic used to determine sample mismatch. However, this small deviation does not appear to reflect systematic sample mismatch and instead could describe transgender and gender nonconforming [14] individuals. We retain these samples with appropriate documentation and encourage researchers utilizing the ATLAS data to perform further sex-based filtering based on their specific analysis criteria.

The final step of genotyping QC involves genotype imputation to the TOPMedFreeze5 reference panel, a multi-ancestry dataset assembled from over 50,000 ancestrally diverse genomes [15]. Ambiguous SNPs were removed as well as SNPs that were not an A, C, G, or T allele. Additionally, indels, duplicate SNPs, and alleles that do not match between the reference panel and the target ATLAS data were removed. Haplotype phasing was performed using Eagle v2.4 [16] and imputation was performed using minimac4 [17] through the Michigan Imputation Server [18]. Both the phasing and imputation steps were performed using the TOPMedFreeze5 reference panel. Overall, approximately 300 million SNPs and indels were used as the backbone for genotype imputation. The imputation process yielded a total of 230 million SNPs from the ATLAS data. We additionally found that SNPs with a lower minor allele frequency (MAF) had lower imputation *r*^2^ scores, demonstrating that rare SNPs were more difficult to accurately impute (Figure 2A) [19–21]. Due to this observation, SNPs with imputation *r*^2^ < 0.90 or MAF < 1% were pruned from the data, leaving a total of 7.9 million well-imputed SNPs across 27,946 individuals for follow-up analyses (Figure 1).

**Figure 2:**
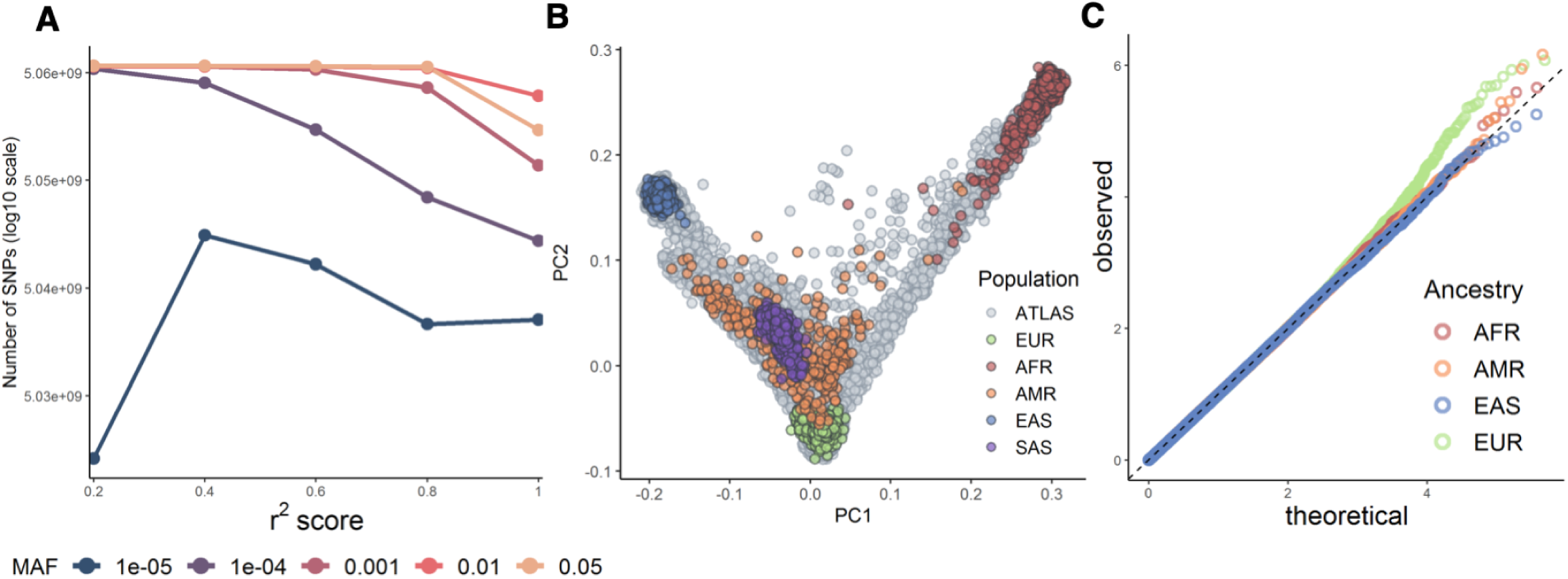
Genotyped and imputed data from ATLAS are of high quality. In A) we show the 7.9 million imputed SNPs binned by the estimated imputation *r*^2^ by quintiles. Within each bin, we show the distribution of SNPs by minor allele frequency (MAF). B) shows the projected genetic PCs 1 and 2 of unrelated individuals in ATLAS (N=27,291) in gray. Samples from 1000 Genomes are shaded by continental genetic ancestry (European, African, Admixed American, East Asian, and South Asian). In C) we show the QQ-plots from the GWAS of gout across the African, Admixed American, East Asian, and European continental ancestry groups within ATLAS.

When performing genome-wide association studies, we stratified individuals by genetic ancestry groups and then performed an additional level of QC separately within each ancestry group. We limited analyses to the subset of 27,291 unrelated individuals (> 2nd degree) and performed ancestry inference (see ‘Genetic ancestry inference’), where each individual was assigned to one continental genetic ancestry cluster: European (N=18,023), African (N=1,340), Admixed American (N=4,930), East Asian (N=2,495), and South Asian ancestry (N=402). At this time, we omitted GWAS analyses within the South Asian ancestry group due to the limited sample size. Individuals who could not be clustered into a specific genetic ancestry group (N=756) were also omitted from GWAS analyses. Within each ancestry group, samples identified as heterozygosity outliers (+/- 3 SDs from the mean) were removed and SNPs that failed the Hardy-Weinberg equilibrium test (*p*-value <1 ×10^−12^) were also removed. Finally, we limited analyses to only SNPs with MAF > 1% within each ancestry group, yielding a total of N=17,874 individuals and M=6.9 million SNPs within the European ancestry group, N=1,337 individuals and M=6.6 million SNPs within the African group, N=4,776 and M=7.2 million SNPs within the Admixed American group, and N=2,459 individuals and M=5.4 million SNPs within the East Asian group.

### Genetic ancestry inference

The ATLAS data presents a unique resource to study genomic medicine across an ancestrally diverse set of individuals within a single medical system. Genetic ancestry information is necessary for numerous types of genetic and epidemiological studies, such as genome-wide association studies and polygenic risk score estimation. The EHR contains self-identified demographic information such as race and ethnicity, but these concepts are distinct from genetic ancestry, which describes the biological history of one’s genome with little to no relation to cultural aspects of identity [22,23]. Although self-identified race/ethnicity and genetic ancestry are correlated [24,25], populations constructed from these two concepts are not analogous and capture distinct information. A thorough discussion of the role of ancestry within the ATLAS data can be found in ref [26].

Instead, we use PCA to identify population structure in ATLAS solely from genetic information as means to correct for genetic stratification in large-scale genotype/phenotype association studies. PCA produces a visual summary of the observed genetic variation which can then be used to describe population structure across the samples. First, we merged genotypes from ATLAS with individuals from the 1000 Genomes Project reference panel [27], which contains genotypes of individuals sampled from various populations: European, African, Admixed American, East Asian, and South Asian. We limited analyses to individuals in ATLAS unrelated to the 2nd degree (N=27,291) and performed PCA analyses on the genotyped data (M=673,130). Genotypes were filtered by Mendel error rate, founders, MAF < 15%, and Hardy-Weinberg equilibrium test (*p*-value < 0.001). Genotypes were then merged with the 1000 Genomes dataset and LD pruning was performed on the merged dataset leaving a total of 253,022 SNPs for the PCA analysis. The top 10 PCs were then computed using the FlashPCA 2.0 software [28].

After projecting the PCs into two-dimensional space, we use the samples from 1000 Genomes to define clusters of individuals corresponding to each continental ancestry group. The first two PCs capture the variation between European, African, and East Asian ancestries. PCs 2 and 3 can approximately delineate individuals with Admixed American ancestry whereas PCs 4 and 5 can cluster individuals with South Asian ancestry (Figure 2B, Supplementary Figure S2). Cluster thresholds were visually determined by comparing the overlap of the 1000 Genomes reference panel samples to ATLAS samples in PC space (Supplementary Figure S3). Individuals who fell into multiple ancestry groups or could not be classified into any of the defined ancestry groups were labeled as ‘Admixed or other ancestry’.

We find that 64.5% (N=18,023) of individuals are inferred to be of European ancestry, 4.8% (N=1,340) of African ancestry, 17.8% (N=4,930) of Admixed American ancestry, 8.9% (N=2,495) of East Asian ancestry, 1.5% (N=402) South Asian ancestry, and 2.7% (N=756) were characterized as ‘Admixed or other ancestry’ (Table 1). As expected, the inferred ancestry clusters were largely concordant with the self-identified race and ethnicity information provided in the EHR: 90.5% of individuals within the European ancestry group self-identified as White/Caucasian, 92.1% of the African ancestry group self-identified as Black or African American, 90.4% of the East Asian ancestry group self-identified as an Asian race, and 77.6% of the Admixed American ancestry group self-identified as either Hispanic or Latino, Puerto Rican, Mexican, or Cuban ethnicity. We observe that most individuals who self-identified to be of African American race tended to fall along the cline between the African and European ancestry clusters, demonstrating that genetic ancestry, in particular for admixed populations, often lies on a continuum rather than within discrete categorizations. These analyses demonstrate how the pairing between self-identified information and inferred genetic ancestry is not one-to-one, further emphasizing the important distinction between these two concepts. A full analysis on exploring the ancestral diversity within ATLAS is described in ref [26].

### EHR-based phenotyping through the phecode system

In this work, we utilized phenotypes derived from the EHR in the form of phecodes, a mapping of ICD codes to a collapsed set of more clinically descriptive groupings [4]. Phecodes allow for systematic phenotyping across a large number of individuals for numerous clinical phenotypes and provide a level of consistency when collaborating across multiple institutions. Additionally, the phecode mapping provides a list of control exclusion phecodes which typically excludes similar or related phecodes to the case phecode. Using both ICD-9 and ICD-10 codes, we constructed 1,866 phecodes using a previously defined ICD-phecode-mapping (Phecode Map 1.2) [29] resulting in a binary phenotype where a patient is a case if the specific phecode occurs at least once within their medical record. Controls are defined as individuals without the occurrence of the case phecode. An additional stricter definition of controls also restricts individuals with the occurrence of any phecode from the case phecode’s control exclusion list.

Out of all individuals in ATLAS (N=27,946), over 99% of individuals have at least one phecode and 30.8% have over 100 distinct phecodes. The distribution of phecodes varies across different demographic groups in ATLAS (Figure 3). Older patients tended to have more phecodes; individuals under the age of 18 had an average of 57.38 (SD: 49.80) unique phecodes and individuals over the age of 64 had an average of 109.98 (SD: 70.34) unique phecodes. We limited subsequent genetic analyses to phecodes with > 100 cases in ATLAS, resulting in a total of 1,330 phecodes.

**Figure 3:**
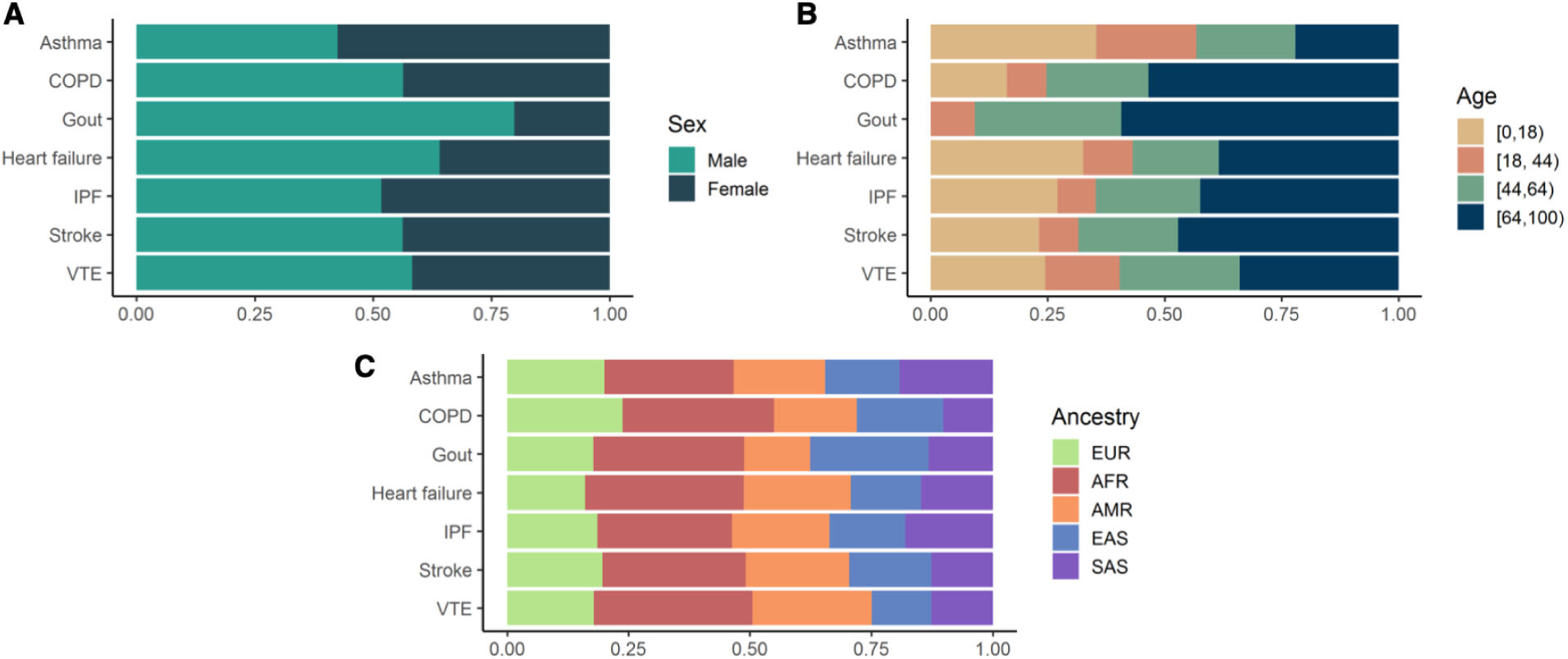
Distribution of phenotypes across different demographic groups in ATLAS. We show the distribution of 7 traits across A) sex, B) age groups, and C) inferred genetic ancestry. See Supplementary Table S1 for the full description of phenotype abbreviations.

To further demonstrate the potential of the EHR-derived phecodes connected with genetic data, we focus on a set of 7 traits to illustrate downstream genetic analyses: asthma, chronic obstructive pulmonary disease (COPD), gout, heart failure (HF), idiopathic pulmonary fibrosis (IPF), cerebral artery occlusion with cerebral infarction (stroke), and venous thromboembolism (VTE). A full list of corresponding phecodes and ICD codes describing these 7 traits is listed in Supplementary Table S1. As shown in Figure 3, the prevalence of certain phecodes varies across sex, age, and genetic ancestry. For example, gout is observed at a much higher frequency in males compared to females (76.4% cases) and tends to be diagnosed in individuals over the age of 64 (59.8% cases). We also observe a high proportion of cases of heart failure within the African ancestry group (freq(all-ATLAS)=0.044, freq(AFR-ATLAS)=0.079; *p*-value=2.4e-6) and cases of gout within the East Asian ancestry group (freq(all-ATLAS)=0.048, freq(EAS-ATLAS)=0.066; *p*-value=8.0e-4) compared to the prevalence across all ATLAS individuals.

### Genome-wide association studies across 7 traits and 4 ancestry groups

As an example of the utility of ancestrally diverse genetic data linked with EHR-based phenotypes (phecodes), we perform GWAS for 7 well-studied traits within each of the 4 continental ancestry groups in ATLAS. The traits represent a wide variety of diseases: asthma, chronic obstructive pulmonary disease (COPD), gout, heart failure (HF), idiopathic pulmonary fibrosis (IPF), cerebral artery occlusion with cerebral infarction (stroke), and venous thromboembolism (VTE) (see ‘EHR-based phenotyping through the phecode system’).

We performed association testing using SAIGE [30], a generalized mixed-model approach that accounts for unbalanced case-control ratios as well as infers and accounts for sample relatedness. Given that many disease phenotypes suffer from case-control imbalance, such as gout (N-case=810, N-control=15,831) and idiopathic pulmonary fibrosis (N-case=700, N-control: 15,941) within the European ancestry group in ATLAS, SAIGE is an advantageous inference method for association testing in ATLAS. In this work, we computed association statistics across

7.3 million SNPs and 27,190 unrelated individuals for 7 traits. Association studies for all 7 traits were performed separately within each of the 4 continental ancestry groups using SAIGE (Table 1) for a total of 28 analyses. Self-identified sex (as reported in the EHR) and current age (as of September 2021) were used as covariates, as well as age*age and age*sex interaction terms. We additionally used the first 10 principal components that were re-computed only on individuals from each ancestry group. Overall, GWAS associations are well-calibrated and do not exhibit strong evidence of test statistic inflation (average across all 28 analyses λ_GC_=0.98, SD(λ_GC_)=0.01) (Figure 2C, Supplementary Figure S4). We found 26 genome-wide significant SNPs (*p*-value < 5 ×10^−8^) within the European ancestry group (gout, heart failure, venous thromboembolism), 1 within the African ancestry group (asthma), and 8 within the Admixed American ancestry group (gout, stroke), for a total of 35 significant SNPs (Figure 4A, Supplementary Table S2).

**Figure 4:**
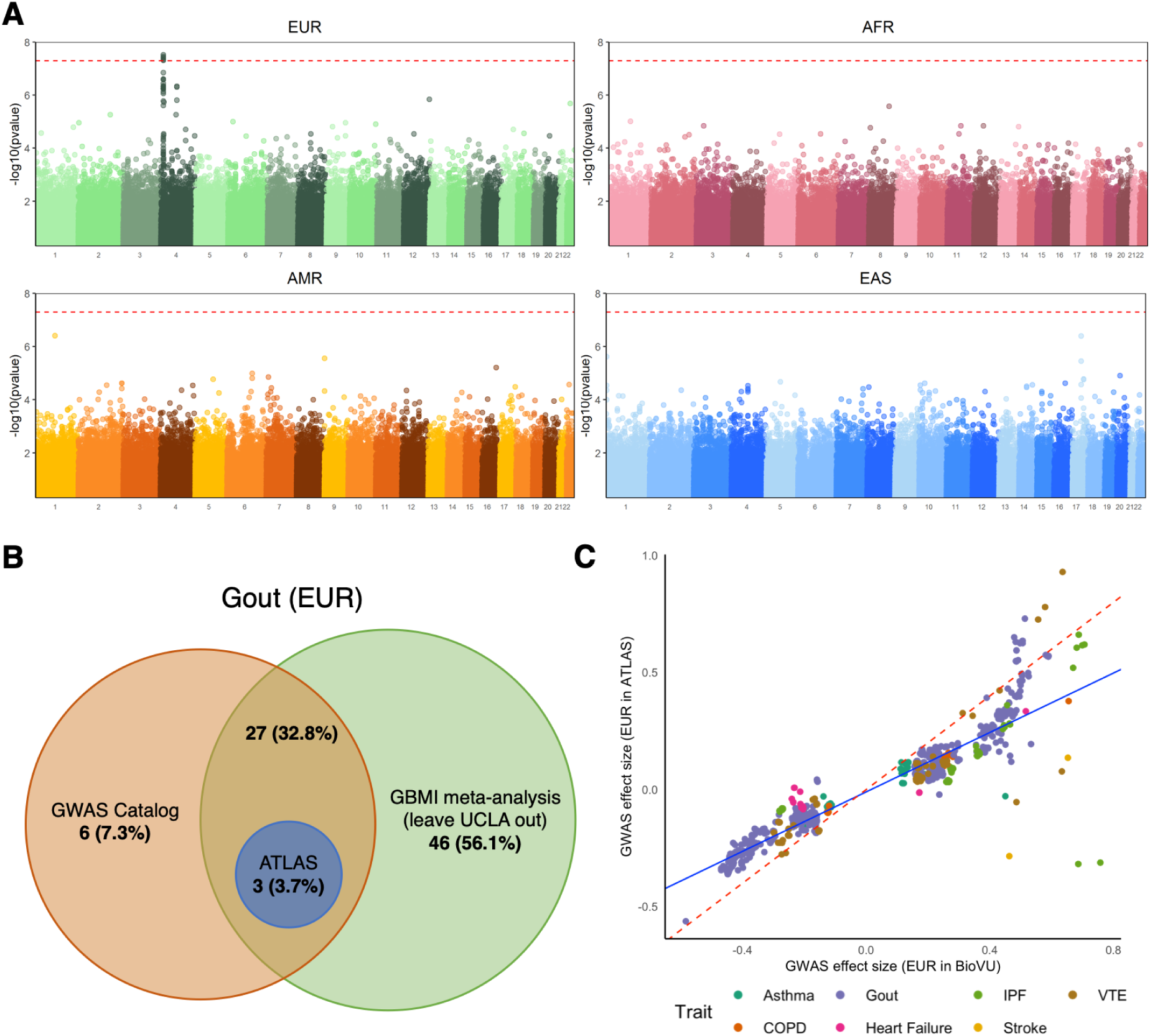
Genome-wide association studies across 7 traits and 4 continental ancestry groups recapitulate previous results. In A) we provide Manhattan plots from the GWAS of gout across the European, African, Admixed American, and East Asian continental ancestry groups in ATLAS. The red dotted line denotes genome-wide significance (*p*-value < 5 ×10^−8^). In B) we show the overlap of genome-wide significant regions for gout computed from ATLAS within the European ancestry group, previous associations listed in the GWAS Catalog, and associations identified in the GBMI meta-analysis. C) shows a scatterplot of GWAS effect sizes of SNPs associated with each trait in either ATLAS and BioVU at *p*-value < 1 ×10^−6^. Points are colored by trait. The red line shows the 45-degree line through the origin, and the blue line shows the estimated trend for these points (Pearson correlation=0.92).

We next compared the associated regions identified in ATLAS to those reported in previous studies, specifically those listed in the GWAS Catalog [31] and the meta-analyses performed through the Global Biobank Meta-analysis Initiative (GBMI) [32]. To avoid biasing our results, we used the GBMI summary statistics that were computed across all other contributing biobanks but omitted ATLAS data from the meta-analysis computation. To construct regions comparable across all of the studies for a given trait, we performed the following procedure. First, we aggregated all SNPs that reached genome-wide significance in at least one of the datasets (i.e. ATLAS, GBMI summary statistics, GWAS Catalog). We then performed a greedy approach by selecting the most significant SNP and created a 1Mb window (500Kb on each side) around this top SNP. All other genome-wide significant SNPs within this window were removed from the list and this procedure was performed until all significant SNPs are accounted for within a region. We defined an individual GWAS for a trait as having a significantly associated region if at least one genome-wide significant SNP fell into one of the constructed regions. Using this process, we found a total of 10 significantly associated regions in ATLAS across the 28 GWAS analyses. Out of these 10 regions, 7 were also reported both in the GWAS Catalog as well as in the GBMI meta-analysis (Figure 4B, Supplementary Figure S5). Finally, when comparing the separate analyses for the 7 traits across the 4 genetic ancestry groups in ATLAS, we did not find any significant associations (SNPs or regions) occurring in multiple populations. This observation could be due to the current limited sample sizes or potentially different genetic architectures across ancestries.

To further assess the congruence of genetic effects estimated in ATLAS to those from more mature EHR-linked biobanks with larger sample sizes, we compared GWAS effect sizes for the 7 traits between ATLAS and BioVU [33] within the European ancestry group. Considering nominally significant SNPs associated with each trait with *p*-value < 1 ×10^−6^ in either study, we find a strong, significant positive correlation (Pearson correlation = 0.92, *p*-value< 2. 2 ×10^−16^) between effect sizes in BioVU and ATLAS (Figure 4C). Although association statistics for the BioVU study were computed using PLINK 2.0 [13] and association statistics for ATLAS were computed using SAIGE, it is encouraging that we observe a positive correlation despite the differences in association testing methods. As shown in Figure 4C, we see that the effects in ATLAS are slightly depressed towards the null, though this may reflect smaller sample sizes in ATLAS compared to BioVU.

### Phenome-wide association studies

EHR-linked biobanks also offer the opportunity to contextualize putative associations within the clinical phenome through phenome-wide association studies (PheWAS) [4] as well as provide a valuable step for validating phenotype quality control. ATLAS has an extensive and diverse set of clinical phenotypes from non-ascertained cohorts which is critical for performing unbiased association tests. We limited our analyses to phecodes with >100 cases within ATLAS, resulting in a total of 1,330 phecodes describing the clinical phenome at UCLA.

To demonstrate the utility of this diverse set of clinical phenotypes, we performed a PheWAS at rs6025, a missense variant within the *F5* gene identified from the ATLAS GWAS of venous thromboembolism in the European ancestry group and also documented in many previous studies [34–36]. We performed an association between rs6025 and 1,330 phecodes and found phenotypic associations with ‘iatrogenic pulmonary embolism and infarction’ and ‘other venous embolism and thrombosis’ (Supplementary Figure S6). Because embolisms generally form in the veins of the leg and then travel to the lungs where they can potentially cause infarctions, these associated phenotypes are consistent with the current understanding of the pathophysiology of venous thromboembolism and pulmonary embolisms [37]. This demonstrates that despite modest sample sizes across many of the phenotypes, we can recapitulate findings consistent with expected disease biology, making PheWAS a valuable tool in investigating the shared genetic architecture across clinical traits.

### Biobank contributions

The ancestral diversity represented in ATLAS plays a key role in the expansion of the catalog of genetic variation used in precision medicine efforts such as polygenic risk scores. Despite its nascency, ATLAS has already contributed to many multi-ancestry disease mapping initiatives, such as the Global Biobank Meta-analysis Initiative (GBMI) [32] and COVID-19 Host Genetics Initiative [38]. Although ATLAS constitutes approximately 1% of the total sample size for the GBMI meta-analysis (N=27,946 samples out of approximately 2.6 million total GBMI samples), we observe a large contribution of samples from diverse ancestral populations within ATLAS to GBMI. For example, ATLAS contributes larger proportions of the African (range of proportions across 7 traits: 3% - 14%) and Admixed American ancestry (22% - 32%) samples when compared to the total sample size in GBMI (Table 2). Additionally, for several phenotypes, ATLAS represents even larger proportions of the total African (AFR) and Admixed American (AMR) ancestry-specific case numbers (e.g., idiopathic pulmonary fibrosis, gout, and heart failure in both AFR and AMR). In addition to GBMI, ATLAS accounted for 73.4% of the Admixed American samples utilized in the primary analysis from the COVID-19 Host Genetics Initiative [38]. This enrichment of AFR and AMR samples from ATLAS can facilitate meta-analytic disease mapping in these historically underrepresented populations and expand the genetic understanding of diverse ancestries.

In the future, we aim to perform phenotyping composed of EHR elements in addition to diagnosis codes, such as laboratory values, medications, and clinical notes. We also plan to incorporate additional types of genomic information such as exome sequencing and methylation data. As sample sizes continue to grow, the ATLAS Community Health Initiative will enable rigorous genetic and epidemiological studies, with a specific aim to accelerate genomic medicine in diverse populations.

## Data Availability

Summary statistic data is contained within the manuscript. All individual-level patient records and genetic data are not available due to privacy.

## Acknowledgments

We gratefully acknowledge the resources provided by the Institute for Precision Health (IPH) and participating UCLA ATLAS Community Health Initiative patients. The UCLA ATLAS Community Health Initiative in collaboration with UCLA ATLAS Precision Health Biobank, is a program of IPH, which directs and supports the biobanking and genotyping of biospecimen samples from participating UCLA patients in collaboration with the David Geffen School of Medicine, UCLA CTSI, and UCLA Health. Additionally, we greatly acknowledge Brett Vanderwerff, Sinéad Chapman, and Benjamin Neale for their insightful feedback as well as all of the members of the Global Biobank Meta-analysis Initiative.

---

**Table 1: Summary of UCLA ATLAS demographics**. We provide summary statistics describing the UCLA ATLAS population computed from data available in the electronic health records and genotype data. Results are computed over all N=27,946 individuals from ATLAS as well as separately within each trait sample size.

**Table 2: UCLA ATLAS contributes a substantial proportion of non-European ancestry samples to global meta-analyses**. We show the case sample sizes across 7 traits for ATLAS and across the entire GBMI study, stratified by genetic ancestry. The last column reports the ratio of the proportion of ancestry-specific samples in ATLAS compared to the proportion of total samples from the GBMI meta-analysis.

## Notes

### Competing Interest Statement

The authors have declared no competing interest.

### Funding Statement

This study did not receive any funding.

### Author Declarations

Patient Recruitment and Sample Collection for Precision Health Activities at UCLA is an approved study by the UCLA Institutional Review Board (UCLA IRB). IRB#17-001013

