## Supplementary Materials for "The UCLA ATLAS Community Health Initiative: promoting precision health research in a diverse biobank"

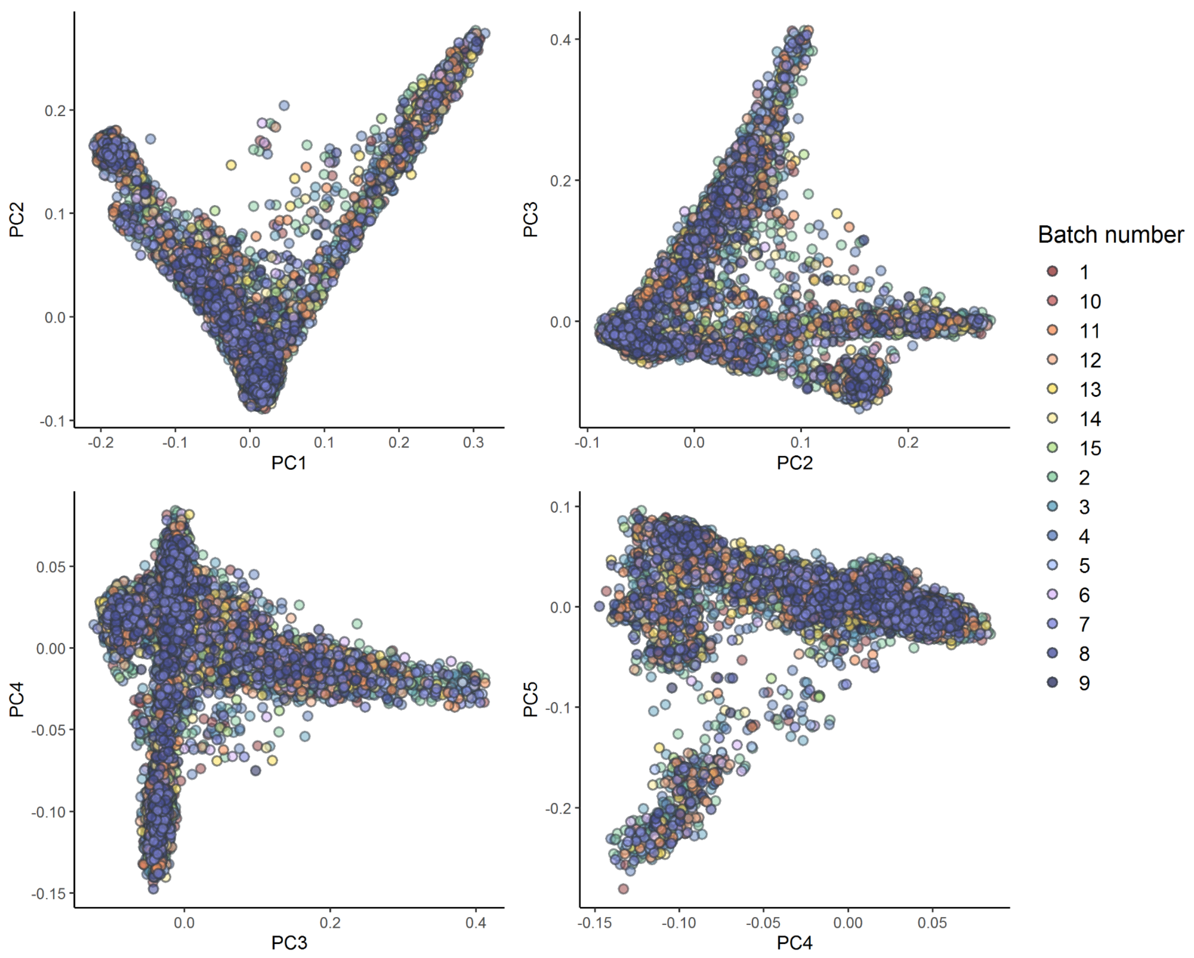


**Supplementary Figure S1: PCA visualization of genotyping batches in ATLAS.** We perform principal component analysis on N=27,291 unrelated ATLAS individuals. We project PCs 1-5 and shade each dot by the sample's batch number.

**Supplementary Figure S2: Summary of genotype quality control pipeline.** We outline the quality control pipeline for the genotype samples and list the number of excluded samples (left) and SNPs (right) at each step.

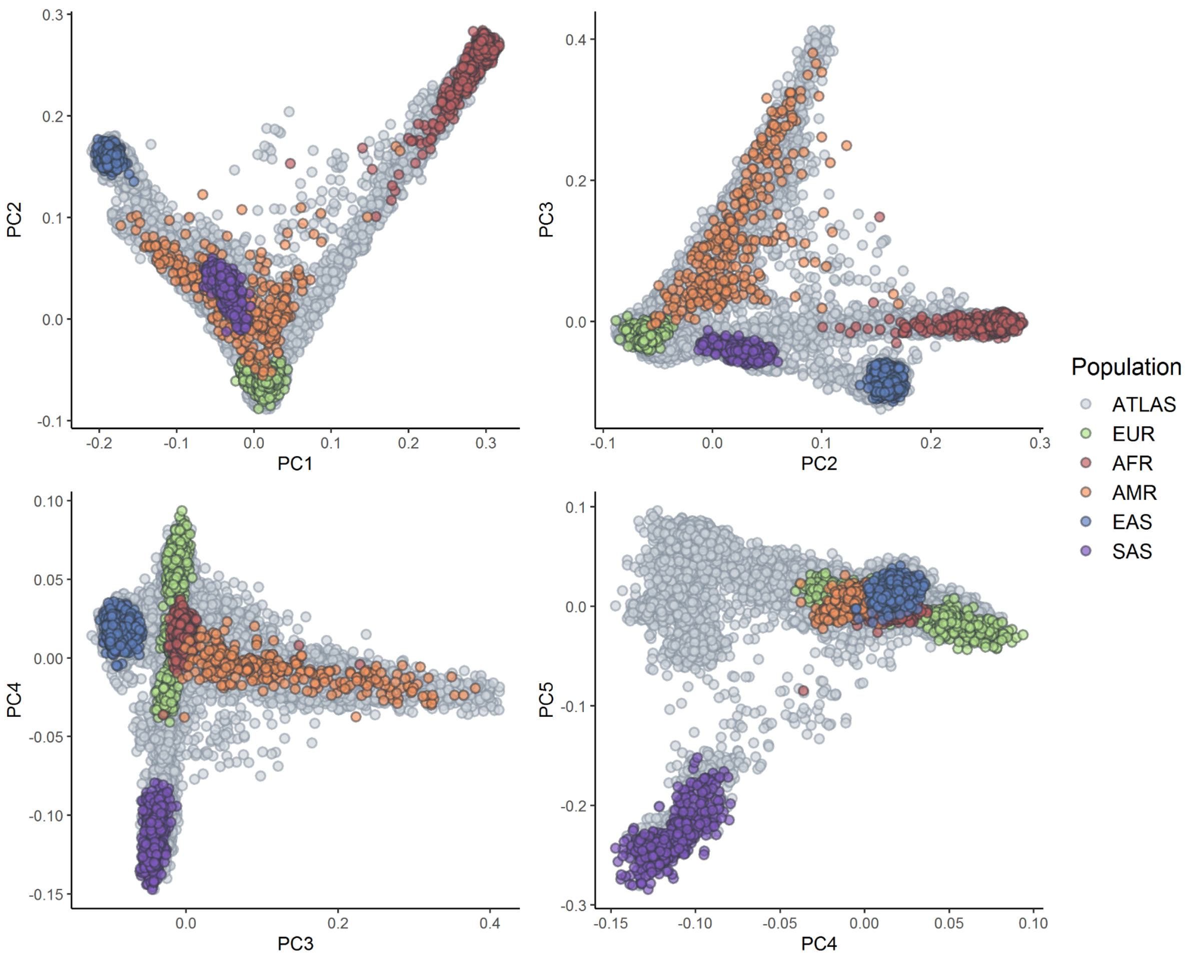


**Supplementary Figure S2: Principal component analysis of ATLAS with 1000 Genomes.** We perform principal component analysis on a merged dataset of unrelated ATLAS individuals (N=27,291) and samples from 1000 Genomes. Individuals from ATLAS are shown in gray and samples from 1000 Genomes are shaded by continental ancestry group.

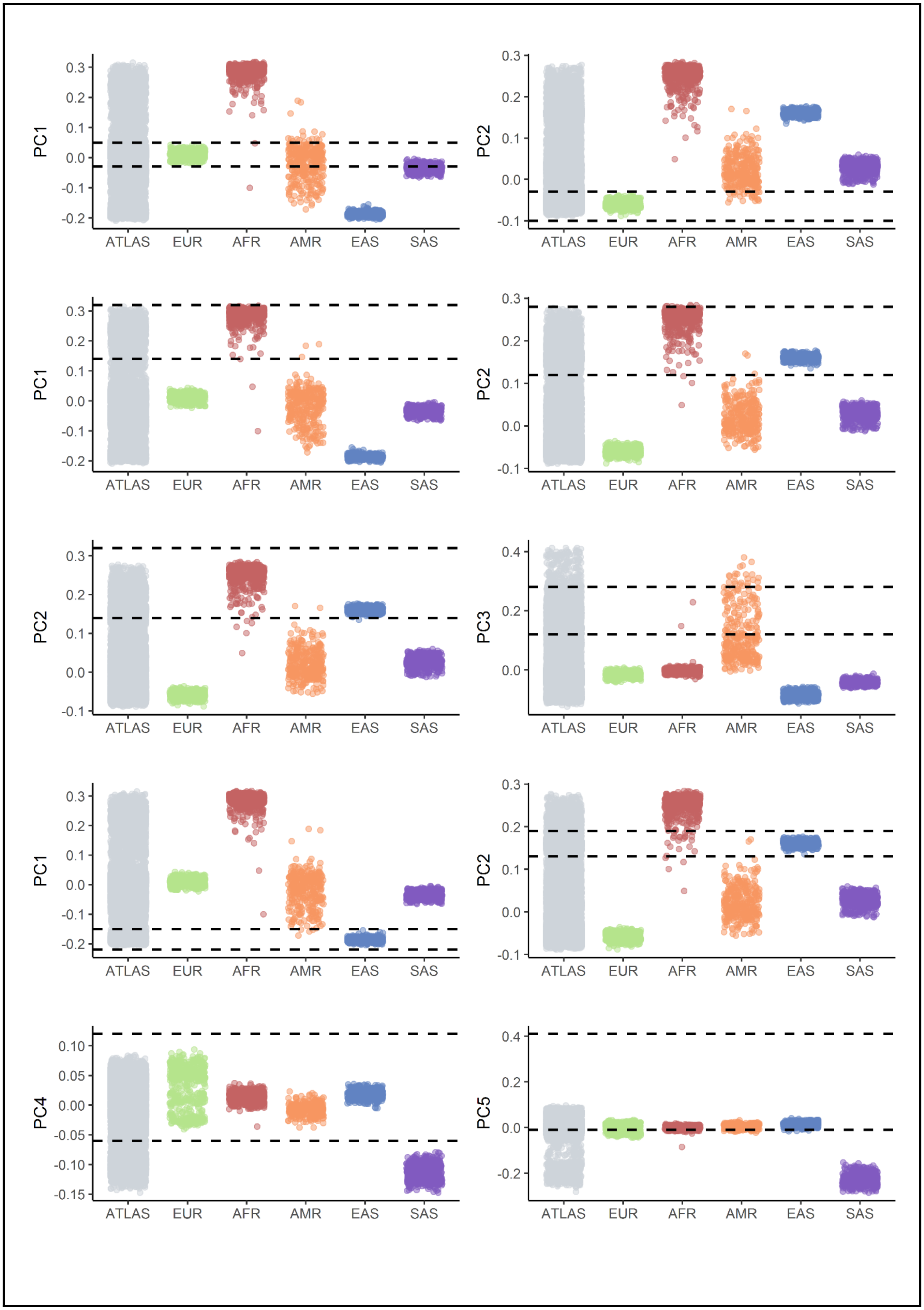
PC1
PC2
PC3
PC4
PC5
ATLAS
EUR
AFR
AMR
EAS
SAS

**Supplementary Figure S3: Inferring continental genetic ancestry through PCA-based clustering.** Genetic PCs of unrelated ATLAS participants (N=27,291) are shown in the first column of each graph in gray and individuals from 1000 Genomes are shaded by continental ancestry. Each continental ancestry cluster is described by two PCs. Dotted horizontal lines denote the thresholds used to define each continental ancestry cluster within ATLAS.

**Supplementary Table S1: (see Excel sheet)**

**Supplementary Table S1: Phecode and ICD codes for 7 EHR-derived phenotypes.** We list the phecodes and corresponding ICD-9 and ICD-10 codes used for defining cases and controls in each GWAS analysis for 7 traits. Phecodes are obtained from phecode mapping v1.2.

**Supplementary Figure S4: (see PDF attachment)**

**Supplementary Figure S4: Summary of GWAS across 7 traits and 4 ancestry groups.** We provide Manhattan plots and QQ-plots for each GWAS analysis. Within the QQ-plot, SNPs that reach genome-wide significance are shaded in dark blue. The red horizontal line in the Manhattan plots denotes genome-wide significance ($p$-value < $5 \times 10^{-8}$). We also provide the sample size, number of SNPs with non-NA $p$-values, the number of SNPs that reach nominal significance ($p$-value < 0.05) and genome-wide significance, and the $\lambda_{GC}$.

**Supplementary Table S2: (see Excel sheet)**

**Supplementary Table S2: List of genome-wide significant SNPs.** We list all SNPs that pass the genome-wide significance threshold ($p$-value$<5 \times 10^{-8}$) for each trait and ancestry group. There were no significant associations identified for any of the traits within the East Asian ancestry group and are therefore not listed in the table. Association studies were not performed within the South Asian ancestry group due to low sample sizes.

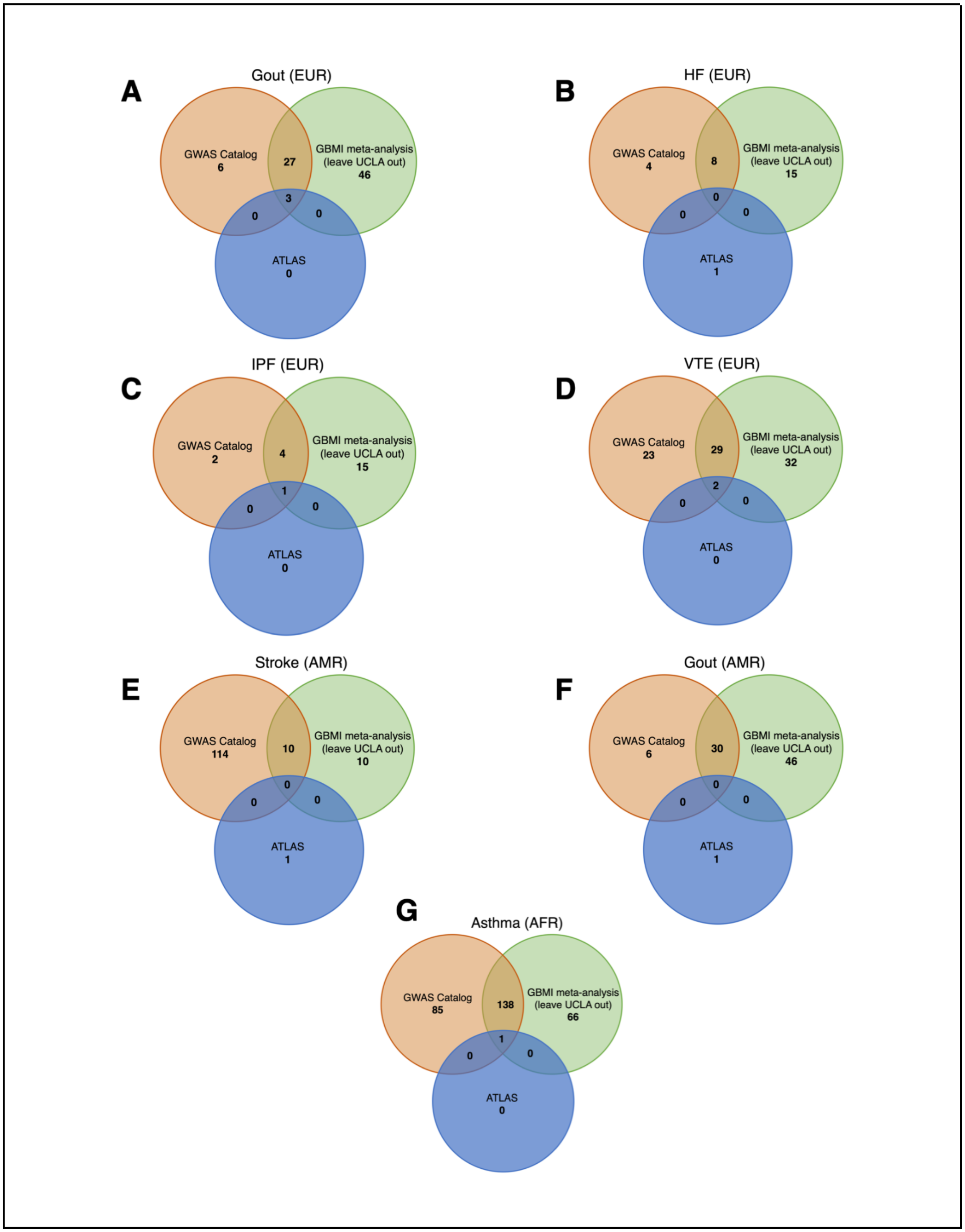
A
Gout (EUR)
GWAS Catalog
6
27
GBMI meta-analysis
(leave UCLA out)
46
3
0
0
ATLAS
0
B
HF (EUR)
GWAS Catalog
4
8
GBMI meta-analysis
(leave UCLA out)
15
0
0
0
ATLAS
1
C
IPF (EUR)
GWAS Catalog
2
4
GBMI meta-analysis
(leave UCLA out)
15
1
0
0
ATLAS
0
D
VTE (EUR)
GWAS Catalog
23
29
GBMI meta-analysis
(leave UCLA out)
32
2
0
0
ATLAS
0
E
Stroke (AMR)
GWAS Catalog
114
10
GBMI meta-analysis
(leave UCLA out)
10
0
0
0
ATLAS
1
F
Gout (AMR)
GWAS Catalog
6
30
GBMI meta-analysis
(leave UCLA out)
46
0
0
0
ATLAS
1
G
Asthma (AFR)
GWAS Catalog
85
138
GBMI meta-analysis
(leave UCLA out)
66
1
0
0
ATLAS
0

**Supplementary Figure S5: Genome-wide significant regions in ATLAS are concordant with previous studies.** We show Venn diagrams that summarize the overlap of genome-wide significant regions from ATLAS, previous associations listed in the GWAS Catalog, and associations identified in the GBMI meta-analysis.

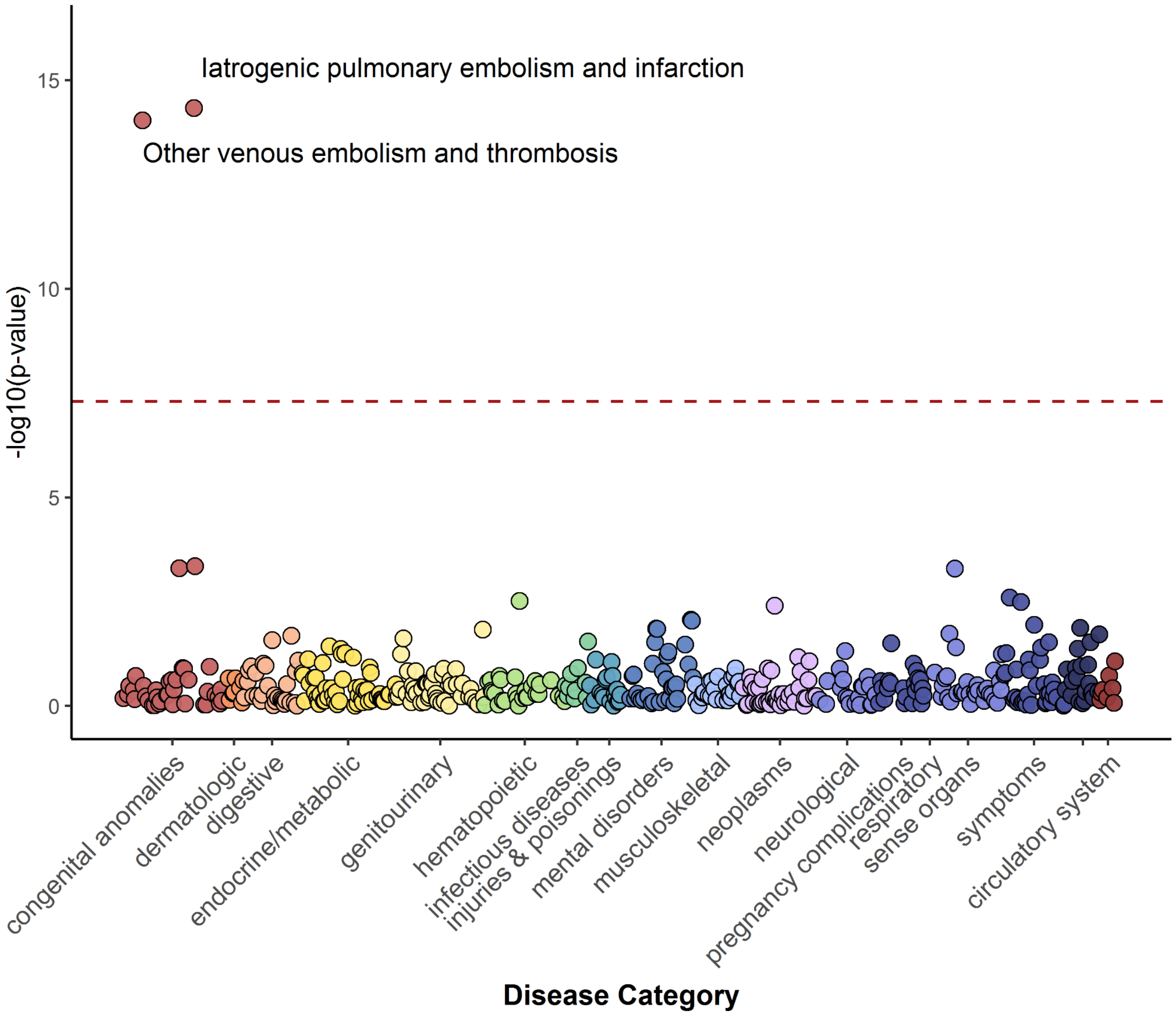


**Supplementary Figure S6: Phenome-wide association study at rs6025.** We show a PheWAS plot at SNP rs6025, a missense variant within the *F5* gene, for the European ancestry group across 1,330 phecodes. The red dotted line denotes $p$-value=$5 \times 10^{-8}$, the genome-wide significance threshold.
