## Supplementary Figure S4 for "The UCLA ATLAS Community Health Initiative: promoting precision health research in a diverse biobank"

**Summary of GWAS across 7 traits and 4 ancestry groups.** We provide Manhattan plots and QQ-plots for each GWAS. The red horizontal line in the Manhattan plots denotes genome-wide significance (*p*-value < 5e-8). SNPs that reach genome-wide significance are shaded in dark blue on the QQ-plot. We also provide the number of SNPs that have non-NA *p*-values, the number of SNPs that reach nominal (*p*-value < 0.05) and genome-wide significance.

### Supplementary Figure S5A: Asthma, EUR

**Cases vs. Controls:** 3051, 13590
**Number of SNPs:** 6859892
**Number of significant SNPs (5e-8):** 0
**Number of nominally significant SNPs (0.05):** 340303
**Lambda GC:** 0.99

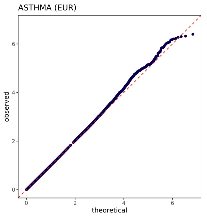


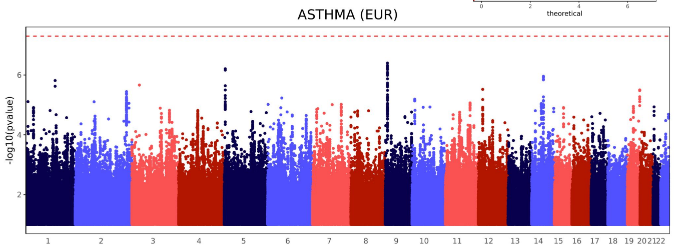


### Supplementary Figure S5B: Asthma, AFR

**Cases vs. Controls:** 289, 937
**Number of SNPs:** 6557250
**Number of significant SNPs (5e-8):** 1
**Number of nominally significant SNPs (0.05):** 319378
**Lambda GC:** 0.99

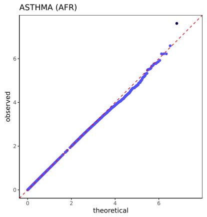


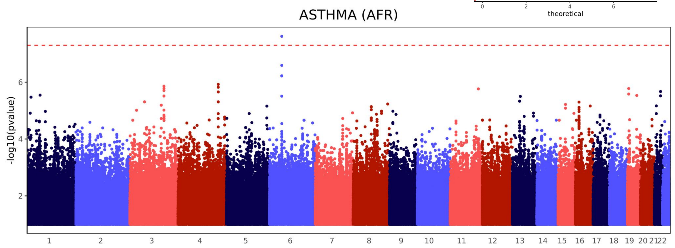


### Supplementary Figure S5C: Asthma, AMR

**Cases vs. Controls**: 760, 3615
**Number of SNPs:** 7055126
**Number of significant SNPs** (5e-8): 0
**Number of nominally significant SNPs** (0.05): 341269
**Lambda GC**: 0.97

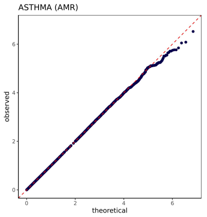


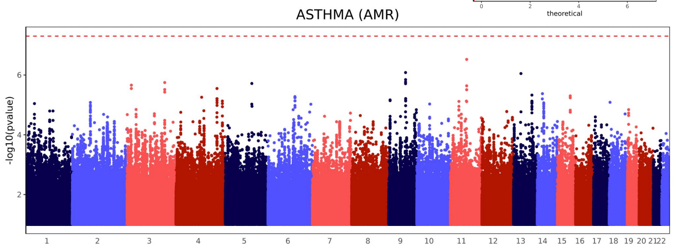


### Supplementary Figure S5D: Asthma, EAS

**Cases vs. Controls:** 308, 1992
**Number of SNPs:** 5396472
**Number of significant SNPs (5e-8):** 0
**Number of nominally significant SNPs (0.05):** 262955
**Lambda GC:** 0.98

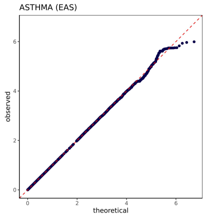


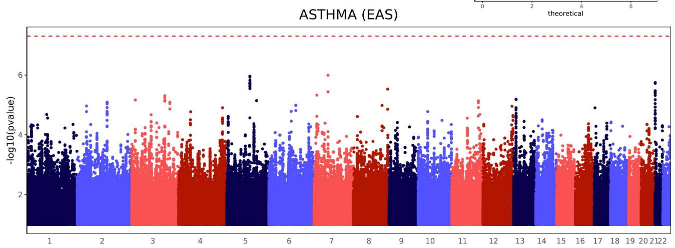


### Supplementary Figure S5E: COPD, EUR

**Cases vs. Controls:** 2005, 14636
**Number of SNPs:** 6850600
**Number of significant SNPs (5e-8):** 0
**Number of nominally significant SNPs (0.05):** 339124
**Lambda GC:** 0.99

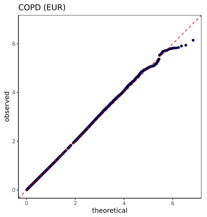


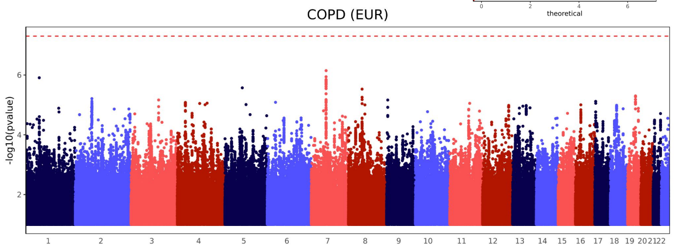

### Supplementary Figure S5F: COPD, AFR

**Cases vs. Controls:** 187, 1039
**Number of SNPs:** 6528611
**Number of significant SNPs (5e-8):** 0
**Number of nominally significant SNPs (0.05):** 315167
**Lambda GC:** 0.98

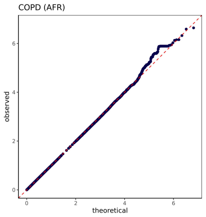


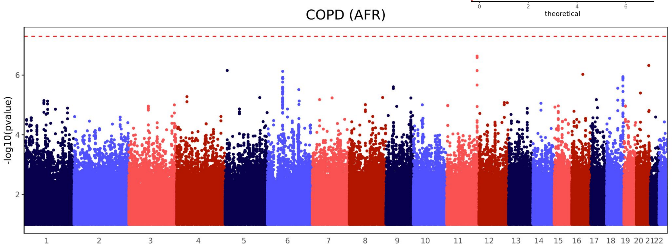


### Supplementary Figure S5G: COPD, AMR

**Cases vs. Controls:** 384, 3991
**Number of SNPs:** 7014983
**Number of significant SNPs (5e-8):** 0
**Number of nominally significant SNPs (0.05):** 334510
**Lambda GC:** 0.96

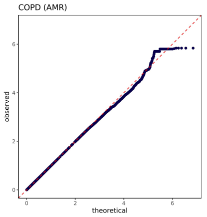


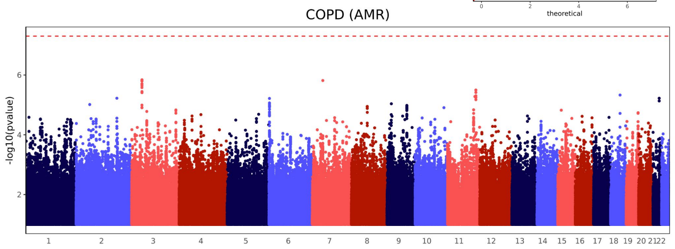


### Supplementary Figure S5H: COPD, EAS

**Cases vs. Controls:** 208, 2092
**Number of SNPs:** 5367966
**Number of significant SNPs (5e-8):** 0
**Number of nominally significant SNPs (0.05):** 260451
**Lambda GC:** 0.99

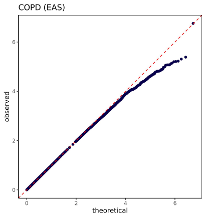


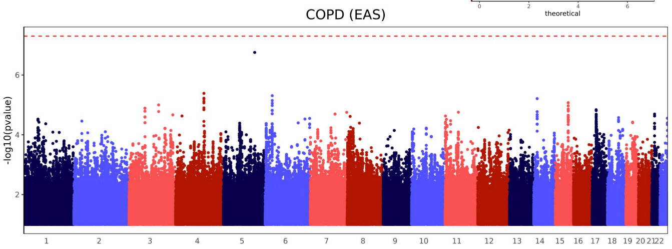


### Supplementary Figure S5I: GOUT, EUR

**Cases vs. Controls:** 810, 15831
**Number of SNPs:** 6824341
**Number of significant SNPs (5e-8):** 20
**Number of nominally significant SNPs (0.05):** 332499
**Lambda GC:** 0.98

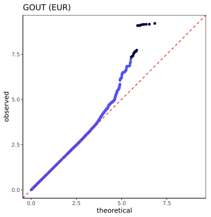


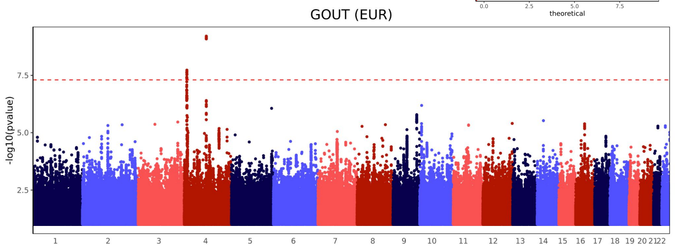


### Supplementary Figure S5J: GOUT, AFR

**Cases vs. Controls:** 105, 1121
**Number of SNPs:** 6428878
**Number of significant SNPs (5e-8):** 0
**Number of nominally significant SNPs (0.05):** 311707
**Lambda GC:** 0.98

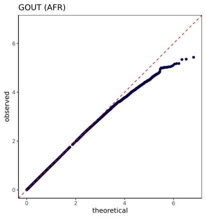


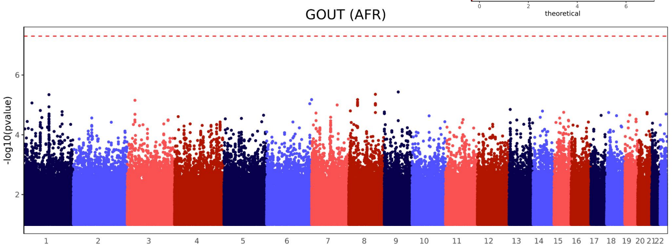


### Supplementary Figure S5K: GOUT, AMR

**Cases vs. Controls:** 179, 4196
**Number of SNPs:** 6946694
**Number of significant SNPs (5e-8):** 3
**Number of nominally significant SNPs (0.05):** 327952
**Lambda GC:** 0.95

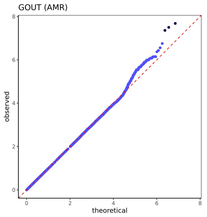


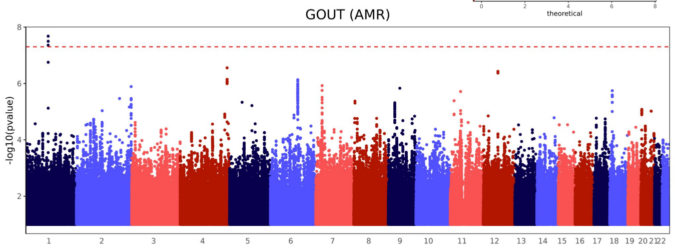

### Supplementary Figure S5L: GOUT, EAS

**Cases vs. Controls:** 155, 2145
**Number of SNPs:** 5353230
**Number of significant SNPs (5e-8):** 0
**Number of nominally significant SNPs (0.05):** 259466
**Lambda GC:** 0.98

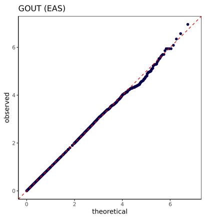


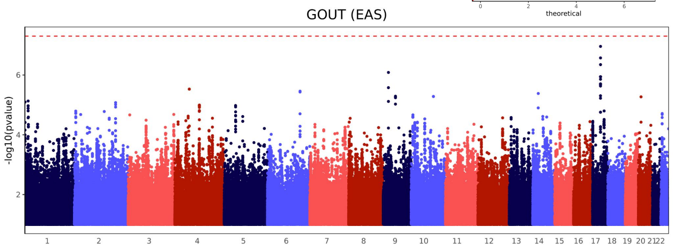


### Supplementary Figure S5M: HF, EUR

**Cases vs. Controls:** 1301, 15340
**Number of SNPs:** 6835981
**Number of significant SNPs (5e-8):** 1
**Number of nominally significant SNPs (0.05):** 331949
**Lambda GC:** 0.99

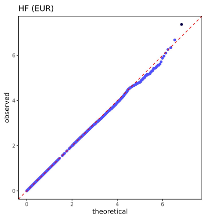


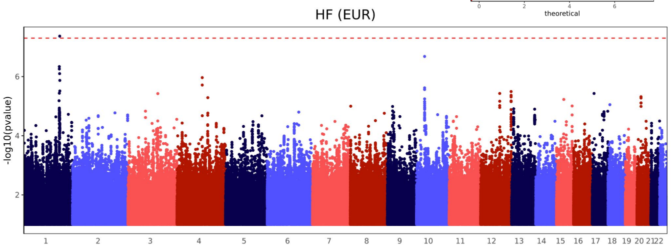


### Supplementary Figure S5N: HF, AFR

**Cases vs. Controls:** 174, 1052
**Number of SNPs:** 6515405
**Number of significant SNPs (5e-8):** 0
**Number of nominally significant SNPs (0.05):** 314043
**Lambda GC:** 0.98

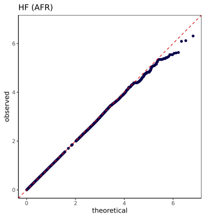


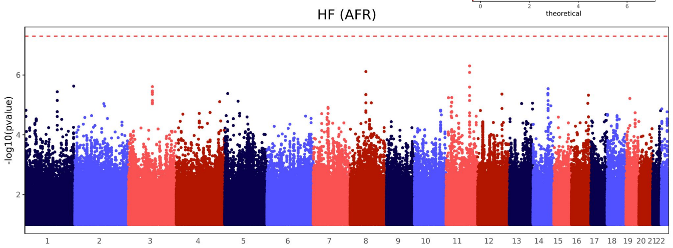


### Supplementary Figure S5O: HF, AMR

**Cases vs. Controls:** 423, 3952
**Number of SNPs:** 6989341
**Number of significant SNPs (5e-8):** 0
**Number of nominally significant SNPs (0.05):** 332637
**Lambda GC:** 0.98

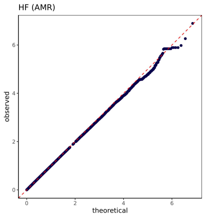


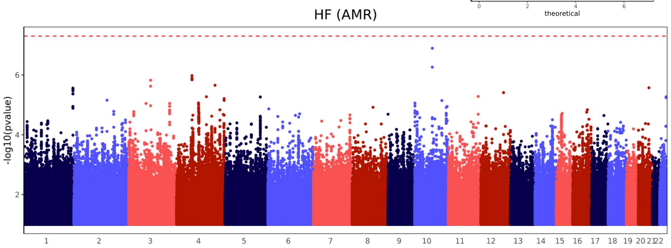


### Supplementary Figure S5P: HF, EAS

**Cases vs. Controls:** 144, 2156
**Number of SNPs:** 5396283
**Number of significant SNPs (5e-8):** 0
**Number of nominally significant SNPs (0.05):** 259905
**Lambda GC:** 0.98

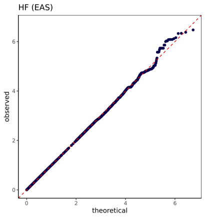


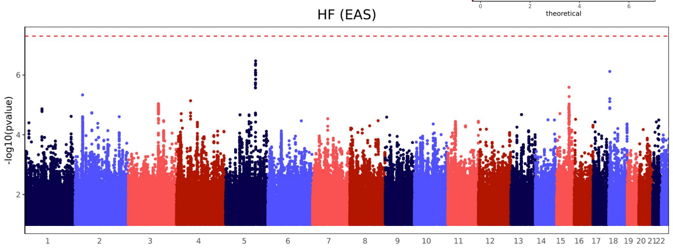


### Supplementary Figure S5Q: IPF, EUR

**Cases vs. Controls:** 700, 15941
**Number of SNPs:** 6825652
**Number of significant SNPs (5e-8):** 2
**Number of nominally significant SNPs (0.05):** 327434
**Lambda GC:** 0.99

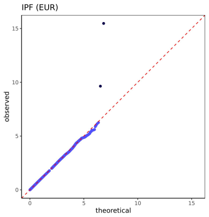


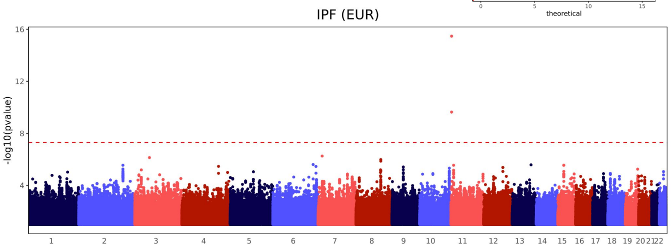

### Supplementary Figure S5R: IPF, AMR

**Cases vs. Controls:** 204, 4171
**Number of SNPs:** 6842352
**Number of significant SNPs (5e-8):** 0
**Number of nominally significant SNPs (0.05):** 323123
**Lambda GC:** 0.96

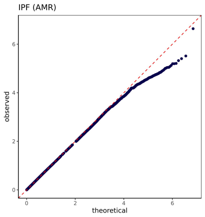


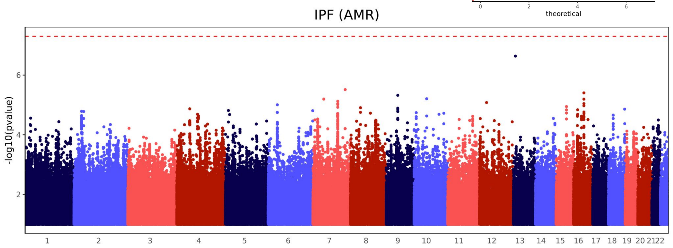


### Supplementary Figure S5S: IPF, AFR

**Cases vs. Controls:** 76, 1150
**Number of SNPs:** 6445498
**Number of significant SNPs (5e-8):** 0
**Number of nominally significant SNPs (0.05):** 306734
**Lambda GC:** 0.96

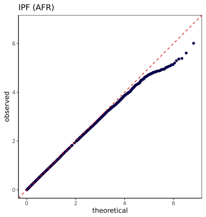


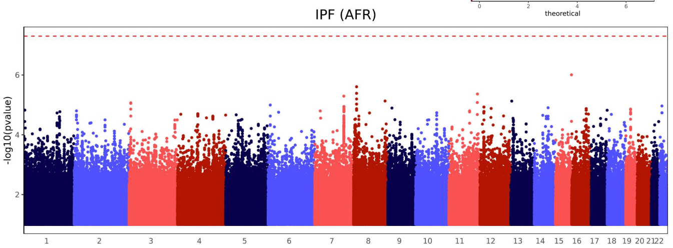


### Supplementary Figure S5T: IPF, EAS

**Cases vs. Controls:** 89, 2211
**Number of SNPs:** 5362117
**Number of significant SNPs (5e-8):** 0
**Number of nominally significant SNPs (0.05):** 253124
**Lambda GC:** 0.97

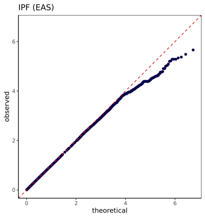


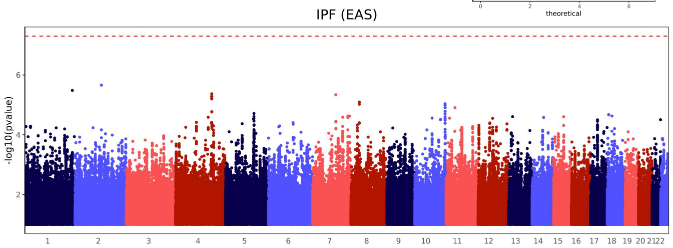


### Supplementary Figure S5U: STROKE, EUR

**Cases vs. Controls:** 855, 15786
**Number of SNPs:** 6821487
**Number of significant SNPs (5e-8):** 0
**Number of nominally significant SNPs (0.05):** 327493
**Lambda GC:** 0.98

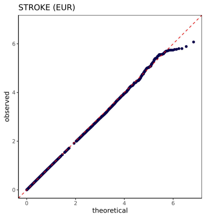


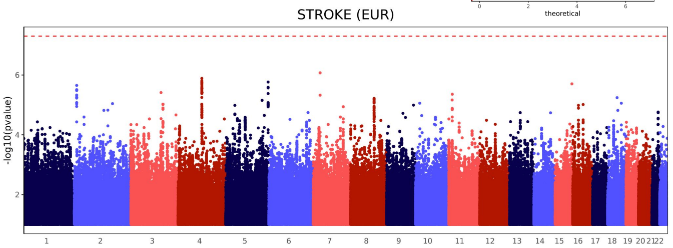


### Supplementary Figure S5V: STROKE, AMR

**Cases vs. Controls:** 248, 4127
**Number of SNPs:** 6989264
**Number of significant SNPs (5e-8):** 5
**Number of nominally significant SNPs (0.05):** 327754
**Lambda GC:** 0.96

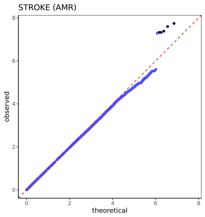


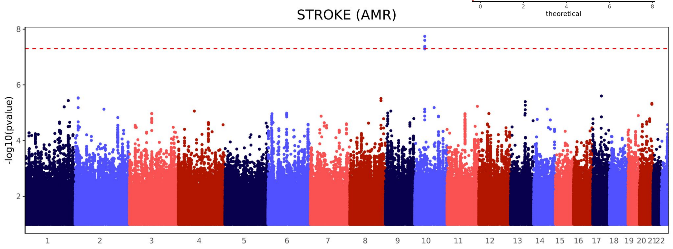


### Supplementary Figure S5W: STROKE, AFR

**Cases vs. Controls:** 100, 1126
**Number of SNPs:** 6515249
**Number of significant SNPs (5e-8):** 0
**Number of nominally significant SNPs (0.05):** 314659
**Lambda GC:** 0.98

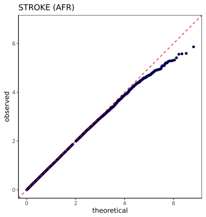


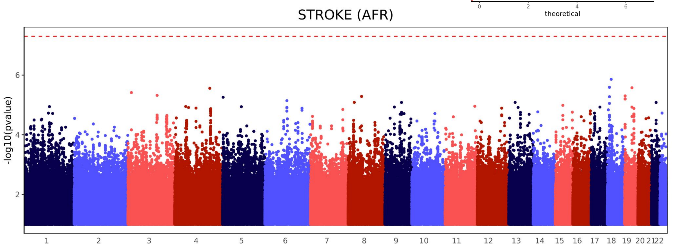

### Supplementary Figure S5X: STROKE, EAS

**Cases vs. Controls:** 105, 2195
**Number of SNPs:** 5312992
**Number of significant SNPs (5e-8):** 0
**Number of nominally significant SNPs (0.05):** 250657
**Lambda GC:** 0.98

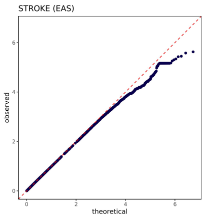


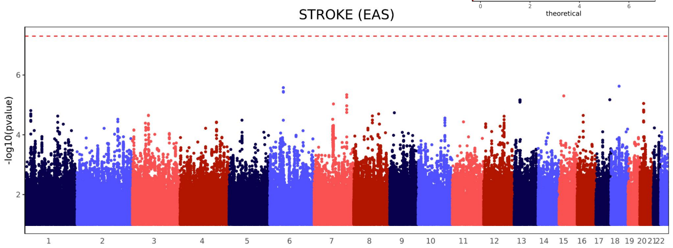


### Supplementary Figure S5Y: VTE, EUR

**Cases vs. Controls:** 1503, 15138
**Number of SNPs:** 6838442
**Number of significant SNPs (5e-8):** 3
**Number of nominally significant SNPs (0.05):** 327404
**Lambda GC:** 0.98

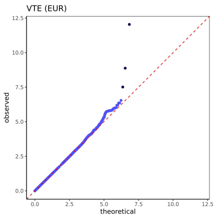


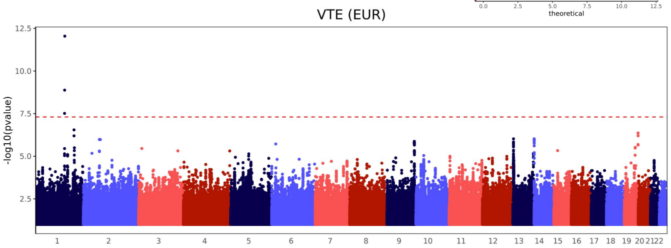


### Supplementary Figure S5Z: VTE, AMR

**Cases vs. Controls:** 543, 3832
**Number of SNPs:** 7032980
**Number of significant SNPs (5e-8):** 0
**Number of nominally significant SNPs (0.05):** 338233
**Lambda GC:** 0.97

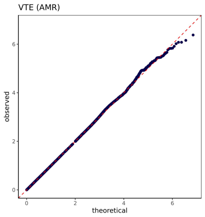


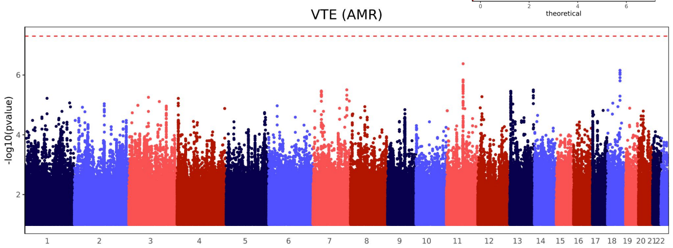


### Supplementary Figure S5AA: VTE, AFR

**Cases vs. Controls:** 195, 1031
**Number of SNPs:** 6534467
**Number of significant SNPs (5e-8):** 0
**Number of nominally significant SNPs (0.05):** 311423
**Lambda GC:** 0.96

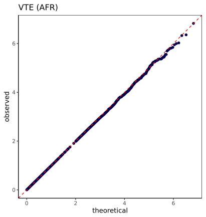


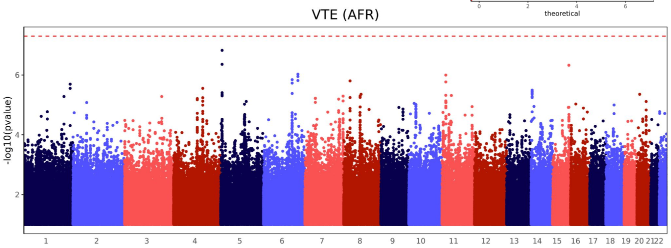


### Supplementary Figure S5AB: VTE, EAS

**Cases vs. Controls:** 132, 2168
**Number of SNPs:** 5373808
**Number of significant SNPs (5e-8):** 0
**Number of nominally significant SNPs (0.05):** 251594
**Lambda GC:** 0.97

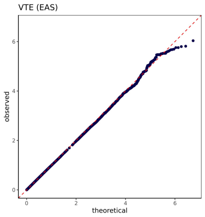


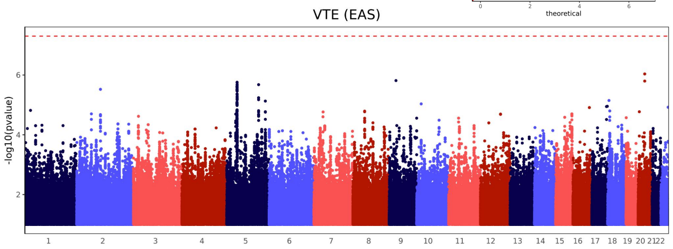
